# The Evaluation of Deep Neural Networks and X-Ray as a Practical Alternative for Diagnosis and Management of COVID-19

**DOI:** 10.1101/2020.05.12.20099481

**Authors:** Mohamed Elgendi, Richard Fletcher, Newton Howard, Carlo Menon, Rabab Ward

## Abstract

High-resolution computed tomography radiology is a critical tool in the diagnosis and management of COVID-19 infection; however, in smaller clinics around the world, there is a shortage of radiologists available to analyze these images. In this paper, we compare the performance of 16 available deep learning algorithms to help identify COVID19. We utilize an already existing diagnostic technology (X-ray) and an already existing neural network (ResNet-50) to diagnose COVID-19. Our approach eliminates the extra time and resources needed to develop new technology and associated algorithm, thus aiding the front-line in the race against the COVID-19 pandemic. Results show that ResNet-50 is the optimal pretrained neural network for the detection of COVID-19, using three different cross-validation ratios, based on training time, accuracy, and network size. We also present a custom visualization of the results that can be used to highlight important visual biomarkers of the disease and disease progression.

## Introduction

On March 11, 2020, the World Health Organization declared the COVID-19 virus as an international pandemic.^1^ The virus spreads between people who are in close contact with one another through respiratory droplets produced by coughing or sneezing.^2^ The current gold standard for COVID-19 detection is real time reverse transcription-polymerase chain reaction (RT-PCR). The test itself takes about 4 hours, however, the process before and after running the test, such as transporting the sample and sending the results, requires a significant amount of time. More importantly, the RT-PCR average turnaround time is 3-6 days, in addition to being relatively costly at an average of CA$4,000 per test^3^. The need for a faster and relatively inexpensive technology for detecting COVID-19 is thus crucial to expedite universal testing.

While the clinical presentation of COVID-19 is very diverse, a critical need is to identify the 20% of patients in which the virus has spread to the type II cells of the alveoli and bronchioles, which lead to hypoxia, organ failure, and death.^4^ In order to meet this need, high-resolution computed tomography (HRCT) X-ray is commonly available worldwide, and the unique pattern of COVID-19 infection progression in the lungs has been identified.^5^ However, despite the widespread availability of X-ray imaging, there is unfortunately a shortage of radiologist in most low-resource clinics and developing countries to help analyze and process these images. For this reason, computer algorithms, such as deep learning, that can automate the process of radiology have begun to attract great interest.^6^ Note that X-ray costs about CA$40 per test^3^, making the use of it as a testing tool more feasible.

Since the initial outbreak of the COVID-19, a few attempts have been made to apply deep learning to detect COVID-19. Narin et al.^7^ reported an accuracy of 98% on a balanced dataset for detecting COVID-19 after investigating three pretrained neural networks. Sethy and Behera^8^ explored 10 different pretrained neural networks, reporting an accuracy of 93% on a balanced dataset, for detecting COVID-19 on X-ray images. Zhang et al.^9^ utilized only one pretrained neural network, scoring 93% on an unbalanced dataset. Hemdan et al.^10^ looked into seven pretrained networks, reporting an accuracy of 90% on a balanced dataset. Apostolopoulos and Bessiana^11^ evaluated five pretrained neural networks, scoring 98% of accuracy on an unbalanced dataset.

However, these attempts did not make clear which existing deep learning method would be the most efficient and robust for COVID-19 compared to many others. Moreover, some of these studies were carried out on unbalanced datasets. Our study aims to determine the optimal learning method, by investigating different types of pretrained networks on a balanced dataset, for COVID-19 testing. Additionally, we attempt to visualize the ResNet-50’s weights, which were used for decision making, on-top of the original X-ray image to visually represent the output of the network.

## Main

To determine the optimal existing pretrained neural network for the detection of COVID-19, we used the CoronaHack -Chest X-Ray-Dataset. The chest X-ray images dataset contains 85 images from patients diagnosed with COVID-19 and 1,576 images from healthy subjects. Five x-ray images collected from the Lateral position were deleted for consistency. Then, we balanced the dataset to include 53 COVID-19 and 53 healthy subjects. After creating a balanced dataset, which is important for producing solid findings, 16 pretrained networks were analyzed following the framework shown in Figure 1.

**Figure 1.**
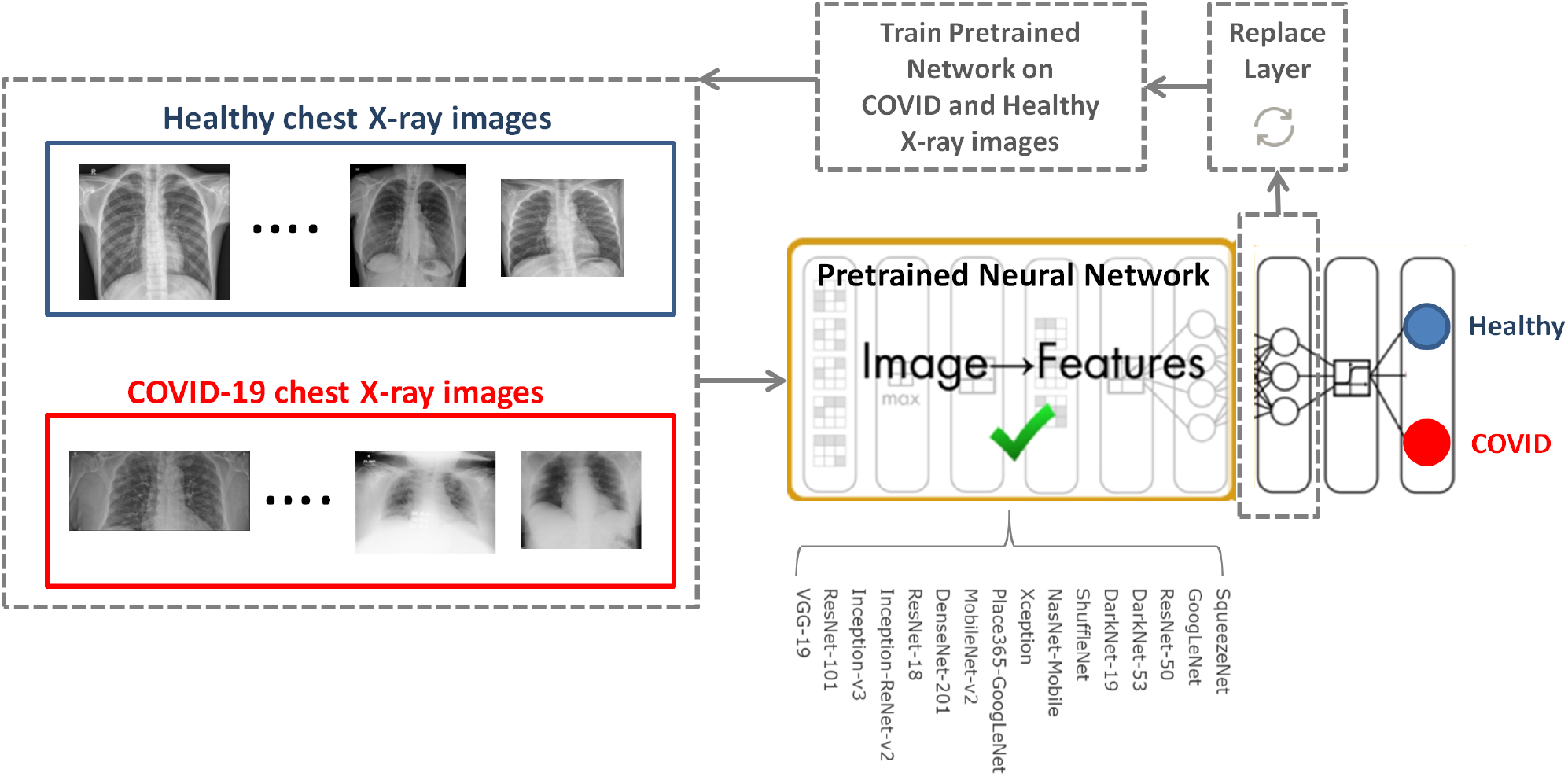
COVID-19 detection framework using pretrained neural networks.

The 16 pretrained neural networks were trained on more than a million images to classify images into 1000 object categories, such as keyboard, mouse, pencil, and many animals. Each network has learned rich feature representations from a large spectrum of images. By replacing the last fully connect layer, as shown in Figure 1, and retrain (fine-tune deeper layers) the neural network with the new dataset (53 COVID-19 and 53 healthy), the neural network can detect COVID-19 and healthy subjects.

The performance of 16 pretrained neural networks using the same dataset (53 COVID-19 and 53 healthy), with different cross validation ratios, is shown in Table1. Interestingly, we found that the following five pretrained neural networks achieved a validation accuracy of 100% using 80-20% cross validation: ResNet-50, DarkNet-53, VGG-19, DenseNet-201, ResNet-18, ResNet-101, and GoogLeNet. The most consistent pretrained neural network in detecting COVID-19, regardless of the cross validation ratio, was ResNet-50, followed by DarkNet-53, followed by VGG-19.

**Table 1.** Performance of 16 pretrained neural networks for detecting COVID-19. The neural networks are ranked in descending order based on the over all performance using three different cross validation ratios.

| Network | 80%-20% CV |  | 70%-30% CV |  | 50%-50% CV |  | Overall Accuracy (%) |
| --- | --- | --- | --- | --- | --- | --- | --- |
|  | Validation Accuracy (%) | Training Time (s) | Validation Accuracy (%) | Training Time (s) | Validation Accuracy (%) | Training Time (s) |  |
| ResNet-50<sup>12</sup> | 100 | 427 | 93.75 | 420 | 98.08 | 296 | 97.28 |
| DarkNet-53<sup>13</sup> | 100 | 763 | 93.75 | 792 | 94.23 | 540 | 95.99 |
| VGG-19<sup>14</sup> | 100 | 1014 | 93.75 | 869 | 92.31 | 678 | 95.35 |
| DenseNet-201<sup>15</sup> | 100 | 889 | 96.88 | 958 | 88.46 | 607 | 95.11 |
| ResNet-18<sup>12</sup> | 100 | 197 | 100 | 224 | 82.69 | 129 | 94.23 |
| DarkNet-19<sup>13</sup> | 95.45 | 349 | 90.63 | 340 | 96.15 | 257 | 94.08 |
| SqueezeNet<sup>16</sup> | 95.45 | 99 | 93.75 | 95 | 92.31 | 76 | 93.84 |
| ResNet-101<sup>12</sup> | 100 | 732 | 78.13 | 623 | 96.15 | 537 | 91.43 |
| GoogLeNet<sup>17</sup> | 100 | 273 | 87.5 | 181 | 84.62 | 134 | 90.71 |
| ShuffleNet<sup>18</sup> | 95.45 | 169 | 84.38 | 163 | 90.38 | 116 | 90.07 |
| MobileNet-v2<sup>19</sup> | 95.45 | 306 | 93.75 | 341 | 75 | 189 | 88.07 |
| Place365-GoogLeNet<sup>17</sup> | 81.82 | 235 | 90.63 | 230 | 84.62 | 125 | 85.69 |
| Inception-v3<sup>20</sup> | 95.45 | 620 | 68.75 | 502 | 76.92 | 429 | 80.37 |
| Xception<sup>21</sup> | 90.91 | 1830 | 65.63 | 1512 | 67.31 | 893 | 74.62 |
| NasNet-Mobile<sup>22</sup> | 81.82 | 702 | 62.5 | 684 | 76.92 | 458 | 73.75 |
| Inception-ResNet-v2<sup>23</sup> | 90.91 | 1187 | 62.5 | 981 | 65.38 | 788 | 72.93 |

DenseNet-201, ResNet-18 and GoogLeNet achieved a validation accuracy below 90% for 50-50% cross validation suggesting that these neural networks are not robust enough for detecting COVID-19 compared to, for example, ResNet-50. In addition, ResNet-101 achieved 87.5% validation accuracy using 70-30% cross validation, suggesting that it is not robust for detecting COVID-19. Despite that the Inception-ReNet-v2 was pretrained on trained on more than a million images from the ImageNet database,^23^ it was ranked the lowest in terms of the overall performance, suggesting it is not suitable to use for detecting COVID-19.

Each pretrained network has a structure that is different from others, e.g., number of layers and size of input. The most important characteristics of a pretrained neural network are as follows: accuracy, speed, and size.^24^ Greater accuracy increases the specificity and sensitivity for COVID-19 detection. Increased speed allows for faster processing. Smaller sized networks can be deployed on systems with less computational resources. Therefore, the optimal network is the network that increases accuracy, utilizes less training time, and that is relatively small in size. Typically, there is a tradeoff between the three characteristics, and not all can be satisfied at once. However, our results show that it is possible to satisfy all three requirements. ResNet-50 outperformed all other networks, while having increased speed and increased accuracy in a relatively small-sized network, as shown in Figure 2. A visual comparison between all investigated pretrained neural networks is presented, with respect to the three characteristics. The x-axis is the training time (logarithmic scale) in seconds, the y-axis is the overall validation accuracy using three different cross validation ratios and the bubble size represents the network size.

**Figure 2.**
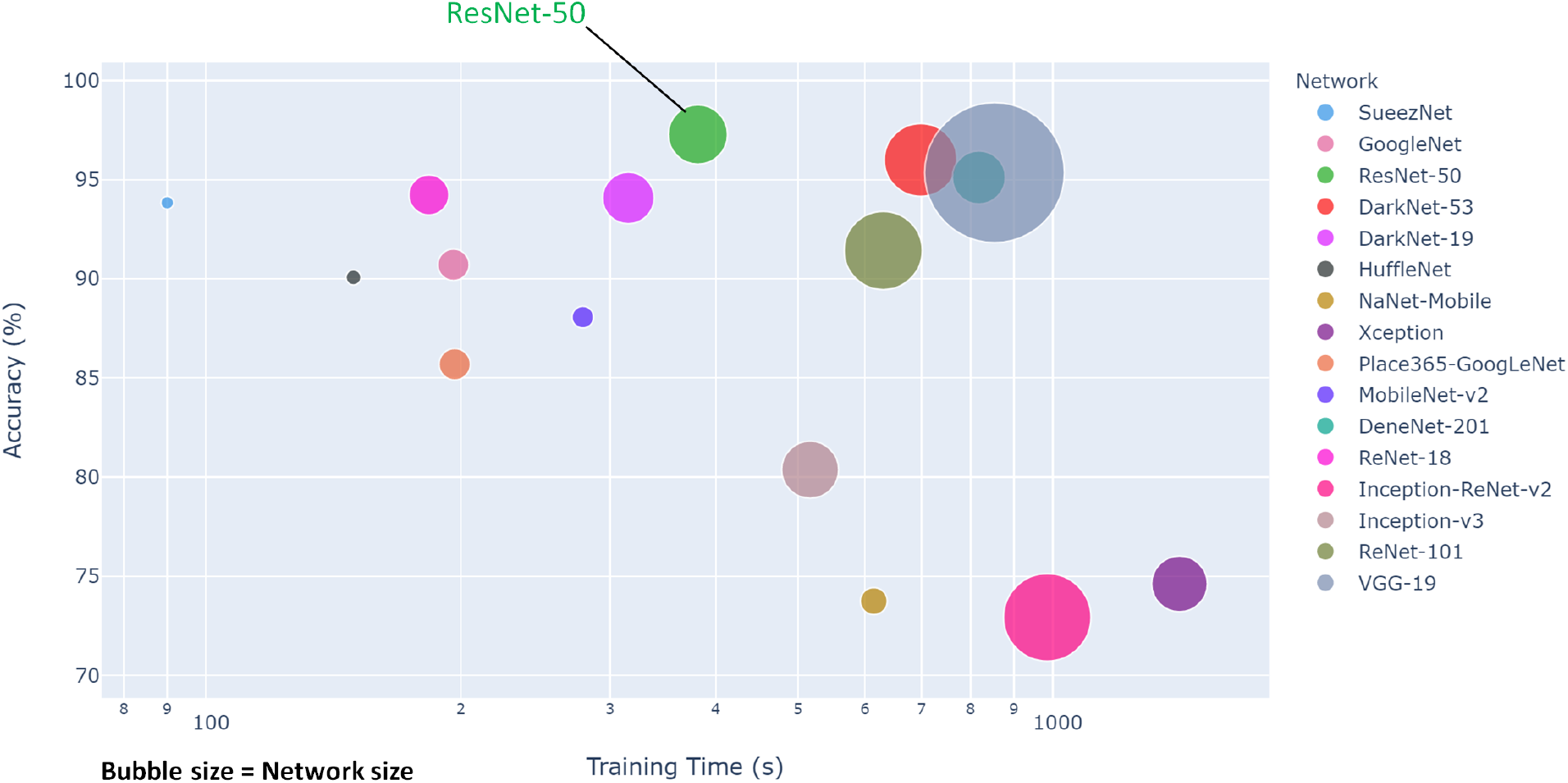
Overall performance of 16 pretrained neural networks for detecting COVID-19.

A comparison of optimal neural networks recommended in previous studies, along with the optimal neural network suggested by this work, is shown in Table 2. Narin et al.^7^ used a balanced sample size of 100 subjects (50 COVID-19 and 50 healthy). They investigated three pretrained neural networks: ResNet50, InceptionV3 and InceptionResNetV2, with a cross validation ratio of 80-20%. They found that ResNet50 outperformed the other two networks, scoring a validation accuracy of 98%.

**Table 2.** Comparison between optimal pretrained neural networks proposed for detecting COVID-19 to date. SVM refers to Support Vector Machine while NC refers to not clear.

| Study | Optimal Network | No. of COVID-19 | No. of Healthy | Cross Validation (%) | Validation Accuracy (%) |
| --- | --- | --- | --- | --- | --- |
| This work | ResNet-50 | 53 | 53 | 80-20 | 100 |
| Narin et al.<sup>7</sup> | ResNet-50 | 50 | 50 | 80-20 | 98 |
| Sethy and Behera<sup>8</sup> | ResNet-50 + SVM | 25 | 25 | 80-20 | 95 |
| Zhang et al.<sup>9</sup> | ResNet-50 | 70 | 30 | NC | 96 |
| Hemdan et al.<sup>10</sup> | VGG19 and DenseNet201 | 25 | 25 | 80-20 | 90 |
| Apostolopoulos and Bessiana<sup>11</sup> | VGG-19 | 224 | 504 | 90-10 | 98.75 |

Sethy and Behera^8^ used a balanced sample size of 50 subjects (25 COVID-19 and 25 healthy). They extracted features from pretrained neural networks and fed them to Support vector Machine (SVM) for classification. They explored the following pretrained neural networks: AlexNet, DenseNet201, GoogleNet, Inceptionv3, ResNet18, ResNet50, ResNet101, VGG16, VGG19, XceptionNet and Inceptionresnetv2, with a cross validation ratio of 80-20%. Again, ResNet50 in combination with SVM outperformed the other networks, with a validation accuracy of 95%.

A similar study by Hemdan et al.^10^ used a balanced sample size of 50 subjects (25 COVID-19 and 25 healthy). The following pretrained neural networks were evaluated: VGG19, DenseNet201, InceptionV3, ResNetV2, InceptionResNetV2, Xception, and MobileNetV2, with a cross validation ratio of 80-20%. Both VGG19 and DenseNet201 scored the same validation accuracy of 90%.

Two studies reported results based on unbalanced datasets: Zhang et al.^9^ and Apostolopoulos and Bessiana^11^. Zhang et al.^9^ created a deep learnig network based on ResNet-50, which achieved an accuracy of 96% with a dataset of 70 COVID-19 and 30 Healthy subjects. Apostolopoulos and Bessiana^11^ used a sample size of 224 COVID-19 and 504 healthy subjects. They tested five pretrained neural networks: VGG19, InceptionV3, InceptionResNetV2, Xception, and MobileNetV2. They found that VGG19 scored highest accuracy of 98.75%, with a cross validation ratio of 90-10%.

Interestingly, ResNet-50 network achieved a validation accuracy of 100%, outperforming other studies that also evaluated ResNet-50 (note that these studies only compared ResNet-50 to a select few neural netowrks, whereas here we compared a total of 16). One possible reason for the difference in performace is that the dataset in our study differed from the datasets in other studies. Another reason is the network’s parameter settings (e.g., learning rate). Note that our finding confirms that ResNet-50 is able to capture COVID-19 from chest X-ray images.

While the ResNet-50 algorithm can distinguish COVID-19 patients from healthy individuals with 100% accuracy, we note the following limitations:

1. The sample size was relatively small, at 58 images for COVID-19 images.
2. The images were not annotated, which is an essential aspect that distinguishes the measurement position of the X-ray image. Thus, we were unable to differentiate between the measurement sites posteroanterior vs. anteroposterior of each X-ray image. Lateral positions were easily identifiable.
3. Our investigation compared COVID-19 patients to healthy individuals. As a next step in our investigation, the X-ray data from COVID-19 patients should also be compared against X-ray data from other respiratory infection patients in order to improve the specificity of the algorithm for detection of COVID-19.

An important component to the automated analysis of the X-ray data is the visualization of the X-ray images, using colors to identify the critical visual biomarkers as well as indication of disease progression. This step can make disease identification more intuitive and easy to understand, especially for healthcare workers with minimal knowledge about COVID-19. The visualization can also expedite the diagnosis process. As shown in Figure 3, COVID-19 and healthy subjects were identified based on the activation images and weights.

**Figure 3.**
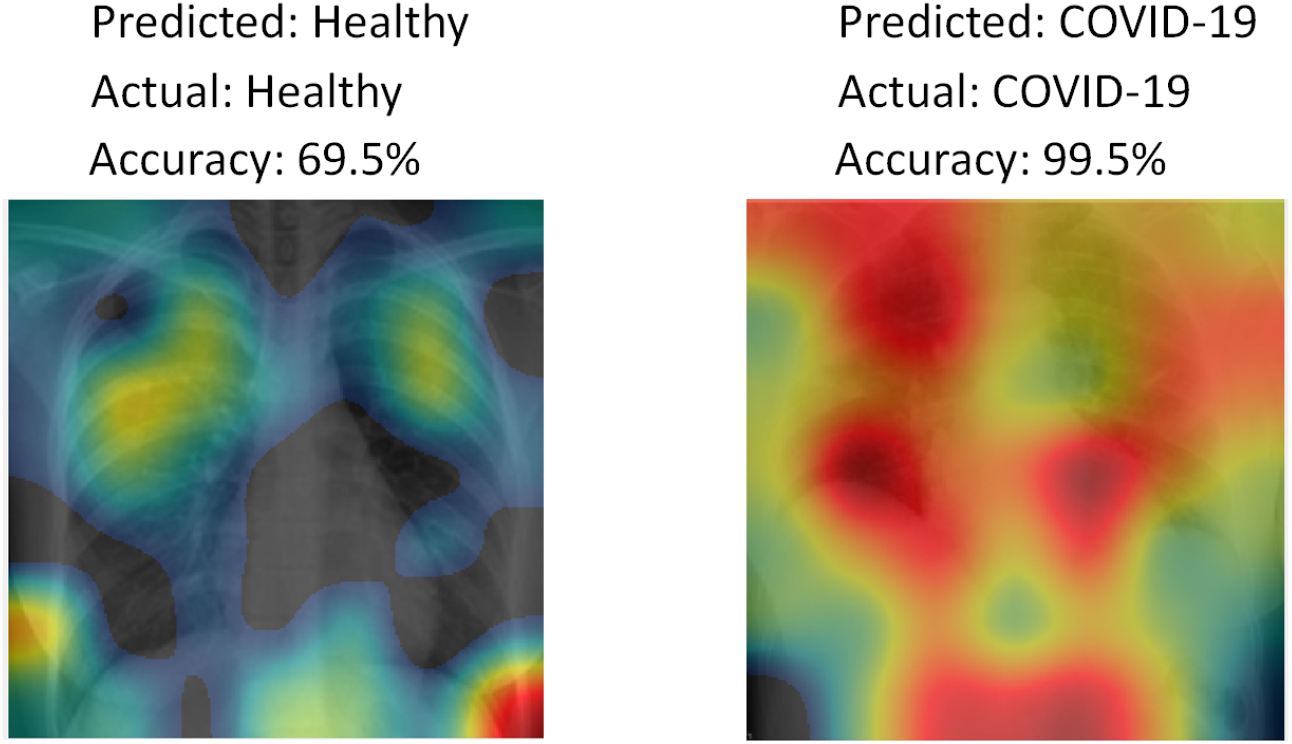
Diagnosing by visualization using class activation mapping.

In conclusion, fast, accurate and accessible tools are needed to help diagnose and manage COVID-19 testing infection. The current gold standard laboratory tests are time consuming and costly, adding delays to the testing process. Chest X-ray imaging for COVID-19 testing is widely available and computer-aided radiology can address existing shortages of radiologists. In this paper, we have reviewed and compared many deep learning techniques currently available in the market for detecting COVID-19 detection in chest X-ray images. After investigating 16 different pretrained neural networks, our results showed that ResNet-50 is the optimal pretrained deep learning network for detection of COVID-19 in HRCT chest X-ray images. Work to improve the specificity of these algorithms in the context of other respiratory infections is ongoing.

## Method

We investigated 16 pretrained neural networks: SqueezeNet^16^, GoogleNet^17^, ResNet-50^12^, DarkNet-53^13^, DarkNet-19^13^, ShuffleNet^18^, NasNet-Mobile^22^, Xception^21^, Place365-GoogLeNet^17^, MobileNet-v2^19^, DenseNet-201^15^, ResNet-18^12^, Inception-ResNet-v28^23^, Inception-v3^20^, ResNet-101^12^, and VGG-19^14^.

All the experiments in our work were carried out in MATLAB 2020a on a PC with the following configuration: 3.70 GHz Intel(R) Core(TM) i7-6500U CPU 2.59 GHz, and 16.00 GB RAM. The dataset was divided into three cross validation sets: 1) 70% training and 30% testing, 2) 80% training and 20% testing, 3) 50% training and 50% testing.

The last fully-connected layer was changed into the new task to classify two classes. The following parameters were fixed for the 16 pretrained neural networks: 1) learning rate was set to 0.0001, validation frequency was set to 5, max epochs was set to 8, and the min batch size was set to 64.

The class activation mapping was carried by multiplying the image activations from the last ReLU layer by the weights of the last fully connected layer of the ResNet-50 network, called “activation_4_9_r_elu,” *as follows*:

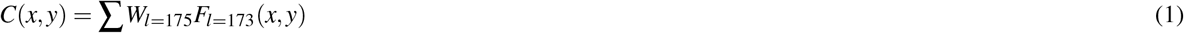

where *C* is the class activation map, *l* is the layer number, *F* is the image activations from ReLu layer (*l* = 173) with dimensions of 7 × 7 × 2048. Here, *W* referes to the weights at *l* = 175 with dimensions of 1 × 2048. Thus, the dimensions of *C* is 7 × 7. Then, we resized *C* to match the size of the original image and visualized it using a jet colormap.

## Data Availability

Publically available dataset

## Data availability

The CoronaHack -ChestX-Ray-Datasetcan be downloadedfrom this link: https://www.kaggle.com/praveengovi/coronahack-chest-xraydataset.

## Author contributions statement

M.E. designed and led the investigation. R.F., N.H., C.M., and R.W. conceived the study and drafted the manuscript. All authors approved the final manuscript.

